# The utility of ChatGPT for cancer treatment information

**DOI:** 10.1101/2023.03.16.23287316

**Authors:** Shan Chen, Benjamin H. Kann, Michael B. Foote, Hugo JWL Aerts, Guergana K. Savova, Raymond H. Mak, Danielle S. Bitterman

## Abstract

The use of large language models (LLMs) such as ChatGPT for medical question-answering is becoming increasingly popular. However, there are concerns that these models may generate and amplify medical misinformation. Because cancer patients frequently seek to educate themselves through online resources, some individuals will likely use ChatGPT to obtain cancer treatment information. This study evaluated the performance and robustness of ChatGPT in providing breast, prostate, and lung cancer treatment recommendations that align with National Comprehensive Cancer Network (NCCN) guidelines. Four prompt templates were created to explore how differences in how the query is posed impacts response. ChatGPT output was scored by 3 oncologists and a 4th oncologist adjudicated in cases of disagreement. ChatGPT provided at least one NCCN-concordant recommendation for 102/104 (98%) prompts. However, 35/102 (34.3%) of these also included a recommendation that was at least partially non-concordant with NCCN guidelines. Responses varied based on prompt type. In conclusion, ChatGPT did not perform well at reliably and robustly providing cancer treatment recommendations. Patients and clinicians should be aware of the limitations of ChatGPT and similar technologies for self-education.

## INTRODUCTION

Large language models (LLMs) underlying chatbots such as ChatGPT^1^ have a unique ability to mimic human language and quickly return detailed and coherent-seeming responses. Yet these properties might obscure the fact that they are providing inaccurate information. Because patients often turn to the internet for self-education,^2^ some will undoubtedly use ChatGPT for cancer-related medical information. This could lead ChatGPT to generate and amplify cancer treatment misinformation. There is thus an immediate need to assess ChatGPT’s performance on these kinds of questions. We evaluated the performance and robustness of ChatGPT to provide breast, prostate, and lung cancer treatment regimen recommendations that are concordant with National Comprehensive Cancer Network (NCCN)^3^ guidelines.

## METHODS

We developed 4 zero-shot prompt templates to query treatment recommendations (Figure). Zero-shot prompts are prompts that do not provide examples of correct responses to guide the model’s output. Templates were used to create 4 prompts for each of 26 unique diagnosis descriptions (cancer types ± extent of disease modifiers relevant for each cancer) for a total of 104 prompts. Prompts were input to the gpt-3.5-turbo-0301 model via the ChatGPT API for inferencing.

**Figure.**
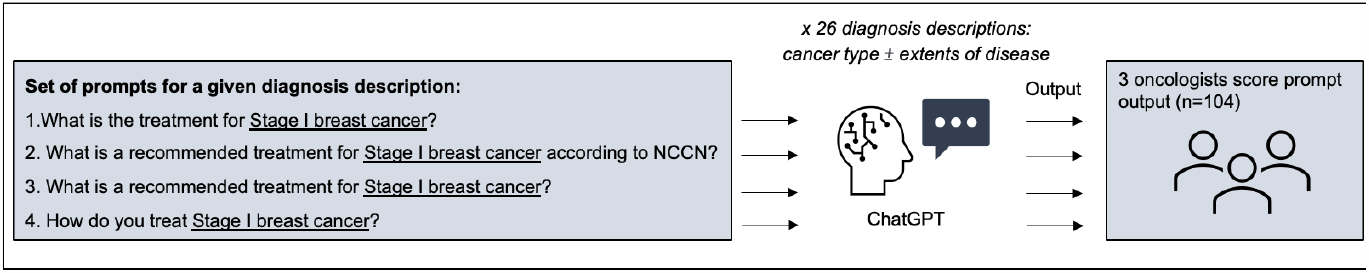
Experimental design. Underlined text indicates where each diagnosis description was input into prompt template. Diagnosis descriptions consisted of cancer type (breast cancer, non-small cell lung cancer, small-cell lung cancer, and prostate cancer) with and without extents of disease relevant for each cancer type. A total of 26 disease descriptions were input into the prompt templates, for a total of 104 unique prompts.

We benchmarked against NCCN 2021 because ChatGPT was trained on data up to September 2021. Five scoring criteria were developed to assess guideline concordance (Table). The output did not have to recommend all possible regimens to be considered concordant; instead, the recommended treatment approach needed to be an NCCN option. Four board-certified oncologists scored output. Prompts were scored by 3 oncologists and majority rule was taken as the final score. In cases of complete disagreement, the oncologist who had not previously seen the output adjudicated.

**Table.** Scoring of ChatGPT treatment recommendations.

| Scoring Criteria | Agreement across 4 prompt templates<sup>a</sup> (n=26) | All prompts (n=104) | Prompts by cancer type |  |  |  |  | Prompts by extent of disease |  |  |
| --- | --- | --- | --- | --- | --- | --- | --- | --- | --- | --- |
|  |  |  | Breast cancer (n=20) | Lung cancer<sup>b</sup> (n=20) | Non-small cell lung cancer (n=20) | Small cell lung cancer (n=12) | Prostate cancer (n=32) | Not Specified (n=20) | Localized (n=64) | Advanced (n=20) |
| 1. How many treatment recommendations are provided? |  |  |  |  |  |  |  |  |  |  |
| None | 22 (84.6%) | 3 (2.9%) | 1 (5.0%) | 1 (5.0%) | 0 | 0 | 1 (3.1%) | 0 | 3 (4.7%) | 0 |
| One |  | 2 (1.9%) | 0 | 0 | 0 | 0 | 2 (6.3%) | 0 | 2 (3.1%) | 0 |
| More than one |  | 99 (95.2%) | 19 (95.0%) | 19 (95.0%) | 20 (100.0%) | 12 (100.0%) | 29 (90.6%) | 20 (100.0%) | 59 (92.2%) | 20 (100.0%) |
| 2. Are the recommended treatments in accordance with NCCN 2021 guidelines?<sup>c</sup> |  |  |  |  |  |  |  |  |  |  |
| None | 9 (34.6%) | 0 | 0 | 0 | 0 | 0 | 0 | 0 | 0 | 0 |
| Some but not all |  | 35 (33.7%) | 2 (10.0%) | 6 (30.0%) | 8 (40.0%) | 7 (58.3%) | 12 (37.5%) | 2 (10.0%) | 26 (40.6%) | 7 (35.0%) |
| All |  | 66 (63.5%) | 17 (85.0%) | 13 (65.0%) | 12 (60.0%) | 5 (41.7%) | 19 (59.4%) | 18 (90.0%) | 35 (54.7%) | 13 (65.0%) |
| N/A |  | 3 (2.9%) | 1 (5.0%) | 1 (5.0%) | 0 | 0 | 1 (3.1%) | 0 | 3 (4.7%) | 0 |
| 3. If "some but not all" to above, are any correct in their entirety per NCCN 2021 guidelines? |  |  |  |  |  |  |  |  |  |  |
| None | 11 (42.3%) | 1 (0.9%) | 0 | 0 | 0 | 1 (8.3%) | 0 | 0 | 1 (1.6%) | 0 |
| At least one |  | 33 (31.7%) | 1 (5.0%) | 7 (35.0%) | 8 (40.0%) | 6 (50.0%) | 11 (34.4%) | 2 (10.0%) | 25 (39.1%) | 6 (30.0%) |
| N/A |  | 70 (67.3%) | 19 (95.0%) | 13 (65.0%) | 12 (60.0%) | 5 (41.7%) | 21 (65.6%) | 18 (90.0%) | 38 (59.4%) | 14 (70.0%) |
| 4. Are any recommended treatments hallucinated? |  |  |  |  |  |  |  |  |  |  |
| None | 16 (61.5%) | 91 (87.5%) | 19 (95.0%) | 18 (90.0%) | 19 (95.0%) | 9 (75.0%) | 26 (81.3%) | 19 (95.0%) | 55 (85.9%) | 17 (85.0%) |
| At least one |  | 13 (12.5%) | 1 (5.0%) | 2 (10.0%) | 1 (5.0%) | 3 (25.0%) | 6 (18.8%) | 1 (5.0%) | 9 (14.1%) | 3 (15.0%) |
| 5. If yes to above, is the hallucinated treatment now a recommended treatment in the most current versions of NCCN |  |  |  |  |  |  |  |  |  |  |
| None | 16 (61.5%) | 13 (12.5%) | 1 (5.0%) | 2 (10.0%) | 1 (5.0%) | 3 (25.0%) | 6 (18.8%) | 1 (5.0%) | 9 (14.1%) | 3 (15.0%) |
| At least one |  | 0 | 0 | 0 | 0 | 0 | 0 | 0 | 0 | 0 |
| N/A |  | 91 (87.5%) | 19 (95.0%) | 18 (90.0%) | 19 (95.0%) | 9 (75.0%) | 26 (81.3%) | 19 (95.0%) | 55 (85.9%) | 17 (85.0%) |

Abbreviations: N/A = Not applicable, NCCN = National Comprehensive Cancer Network.
All results are reported as a N (%), using majority rule of annotators' scores.
<sup>a</sup>Agreement across prompt templates = Percentage of prompts for which the output of each of the 4 prompts for a given diagnosis description yielded the same score.
<sup>b</sup>"Lung cancer" was queried separately to "non-small cell lung cancer" and "small cell lung cancer".
<sup>c</sup>Slight misalignment of categorical scores from Q2 and Q3 sections result from majority rules. For example, Q3 N/A is 70 instead of 69 (66+3) because of majority voting.

All prompts, ChatGPT output, scores, and scoring guidelines are available at https://github.com/AIM-Harvard/ChatGPT_NCCN.

## RESULTS

All 3 annotators agreed on 322/520 (61.9%) scores. The Table shows agreement between prompt templates and the distribution of scores across cancer type and extent of disease. The 4 prompts yielded the same scores for all criteria for 9/26 (34.6%) diagnosis descriptions. ChatGPT provided at least one recommendation for 102/104 (98%) prompts. All outputs with a recommendation included at least one NCCN-concordant treatment, but 35/102 (34.3%) of these outputs also recommended one or more non-concordant treatments.

Modalities were hallucinated (i.e., did not appear as part of any recommended treatment) in 13/104 (12.5%) outputs. These were primarily recommendations for localized treatment for advanced disease, and for targeted therapy or immunotherapy.

## DISCUSSION

One-third of ChatGPT treatment recommendations were at least partially non-concordant with NCCN guidelines, and recommendations varied based on how the question was posed. The disagreement among annotators’ scores highlights the ambiguities and challenges of interpreting generative LLM output—another source of treatment confusion. More work is needed before these methods can be considered for medical question-answering, where both reliability and robustness are critical.

LLMs have been found to achieve a passing grade on the USMLE licensing exam,^4^ encode clinical knowledge^5^, and provide diagnoses better than laypeople.^6^ However, ChatGPT did not perform well at providing cancer treatment recommendations. Concerningly, ChatGPT was most likely to provide incorrect recommendations amongst correct recommendations, an insidious error mode difficult even for experts to detect.

Although this study evaluates a single model at a snapshot in time, it provides insight into areas of concern and future research needs. ChatGPT does not purport to be a medical device, and need not be held to such standards. However, patients and their families will likely use such technologies in their self-education, and this will impact shared decision-making and the patient-clinician relationship.^2^ Developers should have some responsibility to distribute technologies that do not cause harm, and patients and clinicians need to be aware of the limitations of these technologies.

## Data Availability

All data produced are available online at https://github.com/AIM-Harvard/ChatGPT_NCCN

